# On Linear Growth in COVID-19 Cases

**DOI:** 10.1101/2020.06.19.20135640

**Authors:** Michael Grinfeld, Paul A. Mulheran

## Abstract

We present an elementary model of COVID-19 propagation that makes explicit the connection between testing strategies and rates of transmission and the linear growth in new cases observed in many parts of the world. An essential feature of the model is that it captures the population-level response to the infection statistics information provided by governments and other organisations. The conclusions from this model have important implications regarding benefits of wide-spread testing for the presence of the virus, something that deserves greater attention.

## I. INTRODUCTION

Apart from being a world-changing calamity, the present novel coronavirus pandemic is an intellectual challenge for biologists, statisticians and applied mathematicians. Modelling efforts that purport to predict the course of the pandemic and the effect of public health policies, usually take the form of substantial individual-based models and are implemented in code taking thousands of lines. Their predictive ability is disputed but it is doubtless that they do not help us to understand the pandemic. We suggest exactly the opposite: we formulate essentially a two-equation model of one aspect of the pandemic, and claim that it can very simply explain the following puzzling phenomenon: in many countries the rate of appearance of new cases is linear. As an example, we present the data for Sweden in Figure 1 [1].

**FIG. 1.**
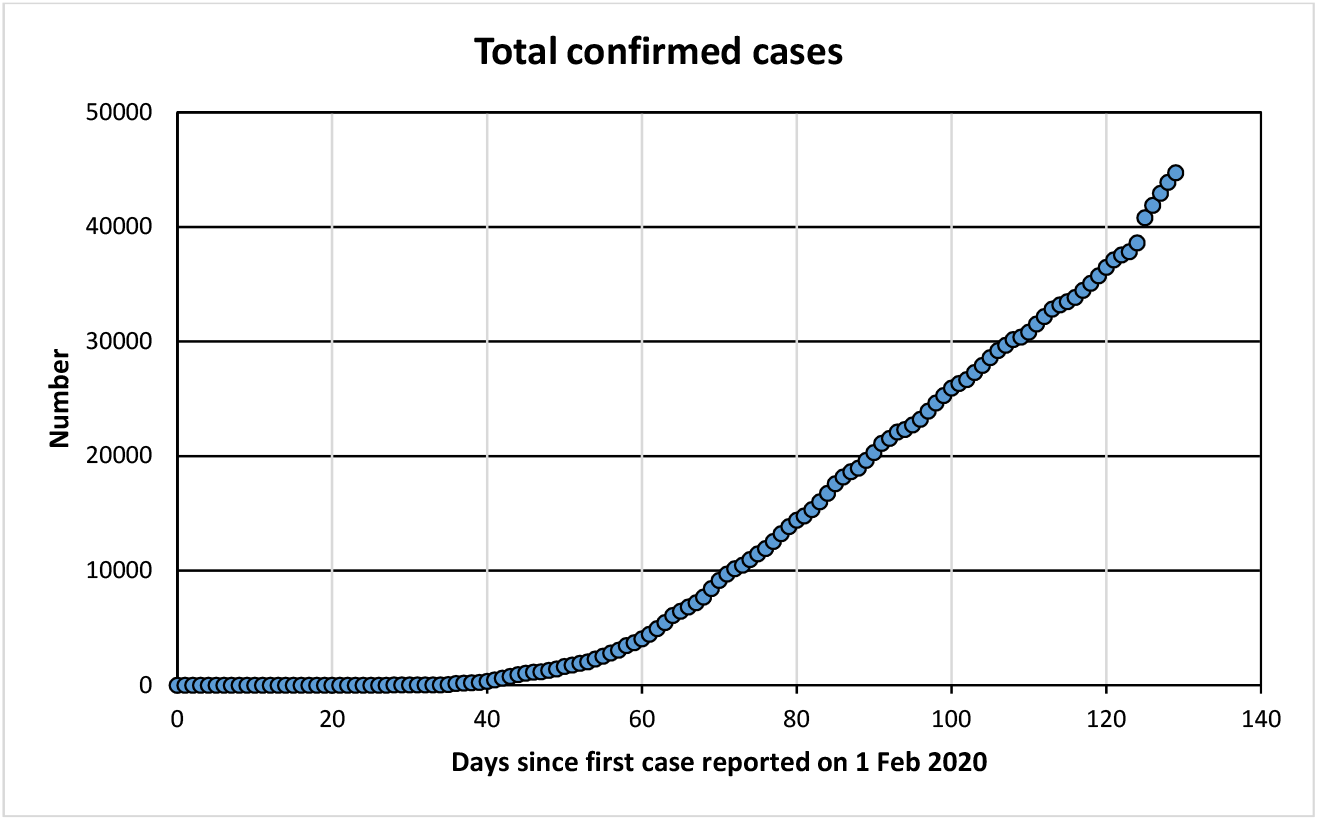
Number of COVID-19 cases in Sweden vs days after 1st case discovered [1].

In fact, Sweden is a good case to work with as there are no complications to do with lock-down; similar graphs can be created, from example, from the data for the state of Georgia [2], among many others.

The modelling of an epidemic on the population level usually divides it in cohorts such as Susceptible, Infective and Recovered (a so-called SIR model), plus possibly some further sub-populations (e.g. Asymptomatic or Exposed (SEIR models)) [3]. The evolution of these cohorts is then modelled using rate equations that include the probability that the disease is transmitted through random contacts between them, amongst other events. In doing this, time is considered as a continuous variable.

However, in order to understand the linear growth phenomenon mentioned above, we believe that it is essential to include the public response to the data that are usually made available on a daily basis. Indeed, we would argue that capturing the response of the population to the information stream is essential if the model is to be of use in truly understanding the pandemic. Therefore, in our approach we will consider time as a discrete variable measured in days, and develop a model for the discrete evolution of the number of infectives from day-to-day. Although unusual, this approach has been successfully used elsewhere in epidemiology; for a recent example, please see [4]

## II. MODELS

We derive, in its simplest and most illuminating form, a system of two difference equations for the rate of growth of new positive test results and the number of people that have been exposed to the virus; that is, we neglect the asymptomatics. The time variable *n* that we use is measured in days. We denote the average latent period (here and below we use epidemiological data from [5, 6]) by *L*; it is about a week. It is known (again, see [5, 6]) that individuals start shedding virus and so are infective very soon (1-2 days) after exposure and about 4-5 days before the appearance of symptoms. Once the simplest model is derived, we consider the case with asymptomatics, which does not offer any substantial new illumination, but is more realistic.

### A. A model without asymptomatics

Let us call the number of positive tests on day *n, T* (*n*).

Then, not taking into account false positives and negatives, but not assuming that every person showing active symptoms is tested,

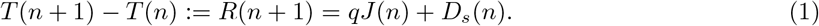

Here *J* (*n*) are people who have shown COVID-19 symptoms on day *n*, and *qJ* (*n*) is the fraction of those who have been tested on that day, and *D*_*s*_(*n*) are the positively testing members of the public who show no symptoms (perhaps yet). The subscript *s* is to indicate that this is only from a sample.

Now, if the rate of testing is *p, D*_*s*_(*n*) = *pD*(*n*), where now *D*(*n*) is the population numbers of people who have virus by PCR but do not show symptoms yet. Let us denote by *E*(*n*) people who got exposed on day *n*. In a model without asymptomatics, *J* (*n*) = *E*(*n − L*) and 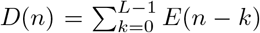 Here *L* is the latent period and for simplicity we have assumed that people become infectious immediately after exposure.

Now we need to model the dynamics of *E*(*n*).

(A1) Since the numbers of infectives are very small compared to the number of susceptibles *S*(*n*), we assume that the number of susceptibles is roughly constant and that *S*(*n*)*/N* ≈ 1, *N* being the total population size.

(A2) We assume that the number of infectives available for infecting the rest of the population on day *n* is approximately *D*(*n*) as *p* is small (of the order of 2*·*. 10^*−*4^ in the UK). Thus, we assume that the moment a person shows symptoms of the disease, she is removed from circulation by hospitalisation or quarantine. That is, we are making an assumption of **perfect isolation**.

It follows from the reasoning above that people who are exposed at time *n, E*(*n*), are determined from the following simple equation:

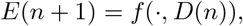

where the other arguments of *f* will be discussed later.

(A3) We assume that *f* (*·*, (*D*(*n*)) can be written as

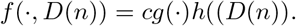

The constant *c* which Hethcote and van den Driessche [7] call “constant contact rate” in our view should only incorporate the probability that an encounter between a susceptible and an infective leads to disease. So presumably wearing face masks or other personal protection measures will be expected to reduce *c*. We take *h*(*D*(*n*)) to be a monotone increasing bounded function with the properties that *h*(0) = 0. e.g. a function of Michaelis-Menten type; see [7] for other examples.

(A4) We assume that the function *g* expresses the information stream of the population, that is, it is the translation of the information that people have into behavioural strategies governing the contact rates of the population (the same function governs also the contact rates between two susceptibles as there is no sure-fire way to determine in a contact between people not showing symptoms who is infected and who is not).

(A5) We assume that the information stream is dominated by the rate of increase of the numbers of new positive tests. It would be interesting to investigate models in which *g* is a function of more than the last day’s data, or of undominated maxima in the number of new cases, but we assume here for simplicity that *R*(*n*) is a reasonable proxy for the information stream. In other words, we assume that *g*(*·*) is a function of *R*(*n*). Common sense suggests that it is a monotone decreasing function defined on ℝ_+_. A possibility is

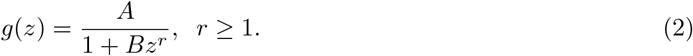

Then *g*(0) = *A* can be interpreted in terms of the norms of sociability in a population. The logic is that if the public is aware of high rate of increase in new cases, it becomes more risk-averse. Note that the information stream is in terms of what is publicly known.

Thus the dynamics of the exposed cohort is governed by

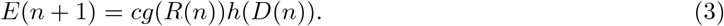

What we have to say about COVID-19 is then summarised in one sentence:

**If the information stream is based on the number of new cases (for which** *R*(*n*) **is a proxy) and quarantining/hospitalisation of symptomatic cases is perfect, linear increase in the number of positive tests is to be expected**.

This is obvious. Clearly from (1), at a fixed point (*R*^*\**^, *E*^*\**^)

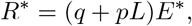

and

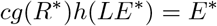

which apart from the fixed point (*R*^*\**^, *E*^*\**^) = (0, 0), in which disease is stopped, may admit a unique non-trivial fixed point, the *E*-component of which solves

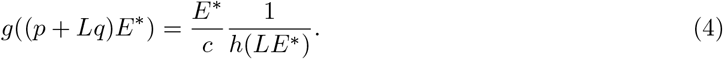

(If such a fixed point does not exist, the epidemic disappears.) If this fixed point is stable, the number of positive tests necessarily grows linearly and from that rate of growth, the number of newly exposed people can be estimated. Note that the value of this equilibrium rate is an increasing function of *c* and also of *L* since the function *g* is monotone decreasing. Under reasonable assumptions on *g* and *h* (for example, *h* being of Michaelis–Menten type and *g* as in (2)) it is easily seen for (4) that *E*^*\**^ is a decreasing function of both *p* and of *q*, since the right-hand side of (4) is monotone increasing and *g* is monotone decreasing. Under these assumptions on *h* and *g*, the equilibrium number of positive tests will grow (sublinearly) with *p* and *q* as is to be expected.

A similar analysis with the same conclusions, can be performed in the case when the information streamis a weighted average 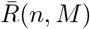 of the rates from a number *M* of days, i.e. if 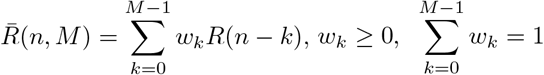; the value of the steady rate is independent on *M* or the weights *w*_*k*_, but these of course influence the stability of the fixed point.

### B. Model with asymptomatics

This model does not add much to our understanding from the previous model, and the purpose of this subsection is simply to show that it subtracts nothing either.

We need to introduce three new parameters: *α* and *β*, and *K. α* and *β* are both in (0, 1). *α* measures the proportion of exposures leading to an asymptomatic state (realistically this seems to be about 0.4 − 0.5) and *β* measures how infectious is an asymptomatic individual relative to an infected one. *K > L* is the average duration of an asymptomatic disease. We need just to modify both (1) and (3)

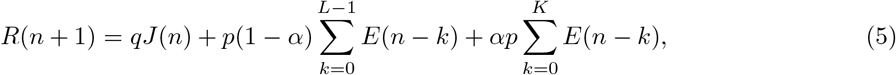

as now the testing also finds some of the asymptomatics who are beyond the latent period. Now *J* (*n*) = (1 *− α*)*E*(*n − L*).

Similarly, (3) becomes

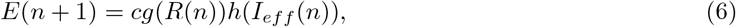

where the number of people available for effective infection on day *n* is

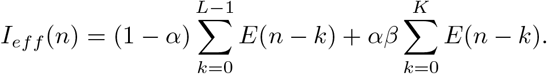

The argument from now is as before. For example, if indeed *α* ≈ 0.5 and as in the UK, *q* ≈ 0.2, with *R*^*\**^ in the UK being approximately 2000, we have that *E*^*\**^ ≈ 20000, i.e. the number of people exposed to the virus each day is about 10 times the number of new cases.

## III. CONCLUSIONS

We found the linear growth rate of the number of COVID-19 positive tests puzzling and have provided a simple framework in which such a dynamic can be expected. In other words, our elementary analysis is in the framework of Peircian abduction, for a good review of which see Psillos [8]. The linear rate is “democratically” determined by the behaviour of the individuals as well as by the rate of testing. We also presented reasons why our assumptions are sensible. We hope the present work is a contribution to the effort to “come to grips” with the pandemic, albeit in a very rough-and-ready and partial fashion; this rough and ready way still allows us, if we have access to additional information, such as that available for the UK from the KCL and ZOE site [9] to estimate the number of asymptomatics, exposed, and effective spreaders.

As a last remark note in the proposed model, government strategy, expressed in the parameters *p* and *q* that determine the published numbers of new cases, directly influences individual behaviour, a feedback loop that does not seem to have been sufficiently discussed. This feedback has to be understood thoroughly in order to craft more effective public health policy.

## Data Availability

All data used is publicly available.

https://en.wikipedia.org/wiki/COVID-19_pandemic_in_Georgia_(U.S._state)

https://ourworldindata.org/coronavirus/country/sweden?country=~SWEphy

https://covid.joinzoe.com/

